# Differential impact of the COVID-19 pandemic on excess mortality and life expectancy loss within the Hispanic population

**DOI:** 10.1101/2023.01.07.23284291

**Authors:** Elizabeth Arias, Betzaida Tejada-Vera

**Affiliations:** National Center for Health Statistics

## Abstract

**Background:** The impact of the COVID-19 pandemic on the Hispanic population resulted in the almost complete elimination of the longstanding Hispanic mortality advantage relative to the non-Hispanic White population. However, it is unknown how COVID-19 mortality affected the diverse Hispanic sub-populations.

**Objective:** We estimate life expectancy at birth in 2019 and 2020 by Hispanic sub-group and explore how changes in age-specific all-cause and COVID-19 mortality affected changes in life expectancy between 2019 and 2020 for each group.

**Methods:** We use final 2019 and 2020 mortality data from the National Center for Health Statistics and population estimates based on the 2019 and 2020 American Community Survey. We calculate life tables and apply decomposition techniques to explore the effects of changes in age- and cause-specific mortality on life expectancy.

**Results:** Patterns of age- and cause-specific excess deaths and their impact on declines in life expectancy due to the COVID-19 pandemic differed substantially by Hispanic sub-group. Life expectancy losses ranged from 0.6 to 6.7 years among males and 0.6 to 3.6 years among females.

**Conclusions:** Our findings highlight the heterogeneous impact of the COVID-19 pandemic within the Hispanic population.

**Contributions:** Our findings contribute new information that will assist future research identify the causes of the disproportionately severe impact of the COVID-19 pandemic on the Hispanic population. Our study underscores the importance of population disaggregation in endeavors to identify the multiple pathways by which the pandemic affected the Hispanic population.

## Introduction

The impact of the COVID-19 pandemic on the Hispanic population has been disproportionate compared to other race and ethnicity groups (Macias Gil et al. 2020; Garcia et al. 2020; Saenz and Garcia 2020; Andrasfay and Goldman 2021; Arias et al. 2022). Until 2020 the Hispanic population experienced lower all-cause mortality than the non-Hispanic White population despite having lower levels of educational attainment, higher levels of poverty, and lower access to health insurance and quality care, a finding first observed over 35 years ago (Markides and Coreil 1986; Markides and Eschbach 2011). In 2019, the Hispanic population had a life expectancy advantage relative to the non-Hispanic White population of over three years (Arias and Xu 2022). By 2021, the advantage had declined to slightly over half a year (Arias et al. 2022). The Hispanic population is a heterogenous grouping of people with origins in numerous countries with distinct cultural, economic, and political systems and who have unique immigration histories and assimilation experiences in the US (Arias et al. 2020). However, it is unknown how the COVID-19 pandemic affected mortality outcomes across the diverse Hispanic origin sub-populations. To our knowledge, there has been only one study that has explored the pandemic’s effect on Hispanic sub-group mortality, but the study is confined to the state of California (Riley et al. 2021).

Life expectancy is a succinct measure of a population’s health status that is easily understood and comparable across populations and time periods. To explore how the pandemic may have differentially affected Hispanic sub-groups, we estimate life expectancy at birth in 2019 and 2020 for the Cuban, Mexican, Puerto Rican, Central and South American (CSA), and Other Hispanic populations. We explore how changes in age-specific all-cause and COVID-19 mortality affected changes in life expectancy between 2019 and 2020 for each sub-group in comparison to the non-Hispanic White population. We restrict the analysis to the first year of the COVID-19 pandemic due to the current unavailability of 2021 population data for Hispanic sub-groups. This limitation does not bias our analysis given that most of the negative effect of the pandemic on Hispanic life expectancy occurred in 2020 (Arias et al. 2022).

## Methods

To estimate life expectancy at birth in 2019 and 2020 for the Hispanic sub-groups, we calculated complete period life tables using the methodology developed by the National Center for Health Statistics (NCHS) to calculate annual life tables for the total Hispanic population. The methodology includes the use of classification ratios developed by NCHS to correct for Hispanic origin misclassification on death certificates and the application of statistical modeling to estimate mortality for the oldest ages (Arias, Heron, and Hakes 2016; Arias and Xu 2022). Classification ratios are only available for the selected sub-groups which prevented the disaggregation of the Central and South American and Other Hispanic sub-groups. The Other Hispanic group includes persons identifying as Dominican, Spaniard, Latin American, Hispanic or Latino/a.

We calculated ratios of the life table age-specific probabilities of dying in 2020 to those in 2019 and used Arriaga’s decomposition method to explore the effects of changes in age and cause-specific mortality on the change in life expectancy between 2019 and 2020 (Arriaga 1984). The age-specific decomposition was done using 5-year age-groups but the results are shown by broad age categories referring to childhood/adolescence (Under 20), working age adults (20-69), and older adults (70 and older).

We used 2019 and 2020 mortality data from NCHS and population estimates based on the full sample of the 1-year 2019 American Community Survey (ACS) and the Public Use Microdata Sample (PUMS) of the 1-year 2020 ACS (U.S. Census Bureau 2019, 2020; National Center for Health Statistics 2019, 2020). Due to the impact of the COVID-19 pandemic, the Census Bureau did not release the standard 1-year ACS estimates for 2020.

## Results

Table 1 presents selected demographic and socioeconomic characteristics of the Hispanic population by country/region of origin. The Hispanic population, the largest minority group, accounted for 18.4% of the total US population in 2019. The Mexican-origin population was the largest, making up 61.5% of the Hispanic population, and the Cuban the smallest (3.9%). Approximately 33% of the Hispanic population was foreign born but there was variation across the sub-groups. Approximately 60% of the Cuban (56.2%) and CSA (59.6%) populations were foreign born in comparison to around 30% of the Mexican, Puerto Rican, and Other Hispanic populations. The Hispanic population was less likely to be 65 and older in comparison to the non-Hispanic White population (20.9%), with the exception of the Cuban population (17.1%).

**Table 1.** Selected demographic and socioeconomic characteristics by Hispanic subgroup in 2019

| **Demographic and Socioeconomic Characteristics** | Hispanic | Cuban | Mexican | Puerto Rican | Central and South American | Other Hispanic | Non-Hispanic White |
| --- | --- | --- | --- | --- | --- | --- | --- |
| Population estimate | 60,481,746 | 2,381,565 | 37,186,361 | 5,828,706 | 9,800,719 | 5,284,395 | 196,789,401 |
| Percent of total population | 18.4 | 0.7 | 11.3 | 1.8 | 3.0 | 1.6 | 60.0 |
| Percent of Hispanic population | 100.0 | 3.9 | 61.5 | 9.6 | 16.2 | 8.7 | ----- |
| Percent foreign born | 32.8 | 56.2 | 29.4 | 28.7 | 59.6 | 30.9 | 4.0 |
| Median age | 29.8 | 40.8 | 28.0 | 30.9 | 32.8 | 31.6 | 43.7 |
| Percent aged 65 and older | 7.7 | 17.1 | 6.5 | 9.5 | 7.4 | 10.4 | 20.9 |
| Percent 25 and older with bachelor's degree or higher | 17.6 | 29.5 | 13.4 | 20.3 | 24.6 | 22.5 | 36.9 |
| Median Household income | \$55,658 | \$56,005 | \$55,943 | \$50,473 | \$59,772 | \$53,712 | \$71,664 |
| Percent without health insurance | 18.7 | 14.0 | 20.3 | 8.0 | 23.4 | 11.8 | 6.3 |
| **Occupation (% civilian employed population 16 and over)** |  |  |  |  |  |  |  |
| Management, business, science, and arts | 23.8 | 33.9 | 21.1 | 29.6 | 25.1 | 26.4 | 44.8 |
| Service | 24.4 | 19.0 | 24.4 | 22.6 | 26.3 | 25.8 | 14.5 |
| Sales and office | 19.5 | 20.8 | 19.3 | 24.0 | 16.8 | 18.2 | 20.8 |
| Natural resources, construction, and maintenance | 15.3 | 10.7 | 17.3 | 8.1 | 16.3 | 14.0 | 8.4 |
| Production, transportation, and material moving | 17.0 | 15.6 | 17.9 | 15.8 | 15.6 | 15.7 | 11.5 |

Notes: Other Hispanic includes Dominicans, Spaniards, and persons identifying as Hispanic/Latino, or Latin American; Occupation titles and their 4-digit codes are based on the 2018 Standard Occupational Classification.
Source: U.S. Census Bureau, 2019 American Community Survey 1-Year Estimates.

All Hispanic sub-groups had lower educational attainment than the non-Hispanic White population, although there was variation. The percentage with a bachelor’s degree or higher ranged from 13.4% for the Mexican population to 29.5% for the Cuban population. All Hispanic subgroups had lower median household income than the non-Hispanic White population. Overall, the Hispanic population had lower percentages of the civilian employed population in higher scioeconomic status occupations (management, business, science and the arts) than the non-Hispanic White population although there were some differences (Fujishiro, Xu and Gong 2010). The percentages in the higher status occupations ranged from 21.1% among the Mexican to 33.9% among the Cuban population. Similarly, the percent without health insurance was higher for all sub-groups in comparison to the non-Hispanic White population although this varied from 8% among Puerto Ricans to 23.4% among CSA.

### Changes in life expectancy at birth

In 2019, the Hispanic population had a life expectancy at birth of 81.9 years (Table 2) (Arias and Xu 2022). Hispanic males and females had life expectancies of 79.1 and 84.4 years, respectively. Within the Hispanic population, life expectancy varied by as much as 6.1 years, with CSAs having the highest (84.4 years) and Other Hispanic the lowest (77.9) life expectancy. Among males, the maximum difference was 7.1 years between CSAs and Other Hispanics. Variation among females was lower at 4.8 years with Cuban females experiencing the highest (86.3 years) and Other Hispanic females the lowest (81.1) life expectancy. Overall, the Hispanic population experienced a life expectancy advantage of 3.1 years relative to the non-Hispanic White population. The advantage varied from 6.5 years for CSAs to -0.9 years for Other Hispanics. Among males, the advantage varied from 7.7 years for CSAs to -1.7 years for Other Hispanics. The advantage varied from -0.2 years for Other Hispanic to 5.0 years for Cuban females.

**Table 2.** Life expectancy at birth by Hispanic subgroup and sex, 2019 and 2020

|  | Life expectancy at birth (Standard Error) |  |  |  |  |  |  |  |  |
| --- | --- | --- | --- | --- | --- | --- | --- | --- | --- |
|  | Total |  |  | Males |  |  | Females |  |  |
|  | 2019 | 2020 | Difference | 2019 | 2020 | Difference | 2019 | 2020 | Difference |
| Hispanic | 81.9 | 77.9 | -4.0 | 79.1 | 74.6 | -4.5 | 84.4 | 81.3 | -3.1 |
| Cuban | 83.6<br>(.113) | 80.5<br>(.099) | -3.1 | 80.7<br>(.169) | 77.7<br>(.145) | -3.0 | 86.3<br>(.137) | 83.2<br>(.116) | -3.1 |
| Mexican | 81.5<br>(.040) | 76.9<br>(.038) | -4.6 | 78.8<br>(.060) | 73.6<br>(.050) | -5.2 | 84.0<br>(.049) | 80.4<br>(.040) | -3.6 |
| Puerto Rican | 80.2<br>(.089) | 76.5<br>(.080) | -3.7 | 77.1<br>(.142) | 73.3<br>(.107) | -3.8 | 83.0<br>(.120) | 79.7<br>(.095) | -3.3 |
| Central & South American | 84.4<br>(.068) | 79.1<br>(.061) | -5.3 | 82.3<br>(.108) | 75.6<br>(.081) | -6.7 | 85.9<br>(.079) | 82.4<br>(.077) | -3.5 |
| Other Hispanic | 77.9<br>(.099) | 77.3<br>(.062) | -0.6 | 74.6<br>(.157) | 74.0<br>(.091) | -0.6 | 81.1<br>(.119) | 80.5<br>(.081) | -0.6 |
| Non-Hispanic white | 78.8 | 77.4 | -1.4 | 76.3 | 74.8 | -1.5 | 81.3 | 80.1 | -1.2 |

Notes: Other Hispanic includes Dominicans, Spaniards, and persons identifying as Hispanic/Latino, or Latin American.
Source: National Center for Health Statistics, National Vital Statistics System, Mortality.

Life expectancy declined by 4.0 years for the Hispanic population in 2020 to 77.9 years (Table 1) (Arias and Xu 2022). Decreases in life expectancy varied significantly by sub-group. The CSA population experienced the greatest decline (−5.3 years) and Other Hispanic the smallest (−0.6). Hispanic males lost 4.5 years of life expectancy between 2019 and 2020 with losses varying from 6.7 to 0.6 years for CSAs and Other Hispanic, respectively. Hispanic females lost 3.1 years, ranging across subgroups from 3.6 years for Mexican and 0.6 years for Other Hispanic populations. In 2020, the Hispanic mortality advantage was reduced to 0.5 years. It declined to 1.2 years for females and became a disadvantage of -0.2 years for males. Across the subgroups, only Cuban and CSA populations retained an advantage relative to the non-Hispanic White population in 2020 (3.1 and 1.7, respectively). Similarly, among males, only Cuban and CSA populations experienced advantages (2.9 and 0.8, respectively). Cuban, CSA, and Mexican females retained advantages of 3.1, 2.3, and 0.3, respectively.

### Changes in age-specific mortality

Figures 1a-1b show ratios of the age-specific probabilities of dying in 2020 to those in 2019 for the non-Hispanic White, Hispanic, and Hispanic sub-groups by sex for ages 20 to 100+. Due to very small numbers of deaths, ratios for ages under 20 vary widely and thus are not shown. The ratios show the large disparities in relative increases in mortality between the non-Hispanic White and the Hispanic population. The age patterns of increases in mortality also differed between the two populations, especially among males. While the age pattern of mortality increases was relatively stable for non-Hispanic White males, for example, the pattern for Hispanic males demonstrated large increases and decreases with age with the greatest increases in mortality in the age-range 35 through 70.

**Figure 1a:**
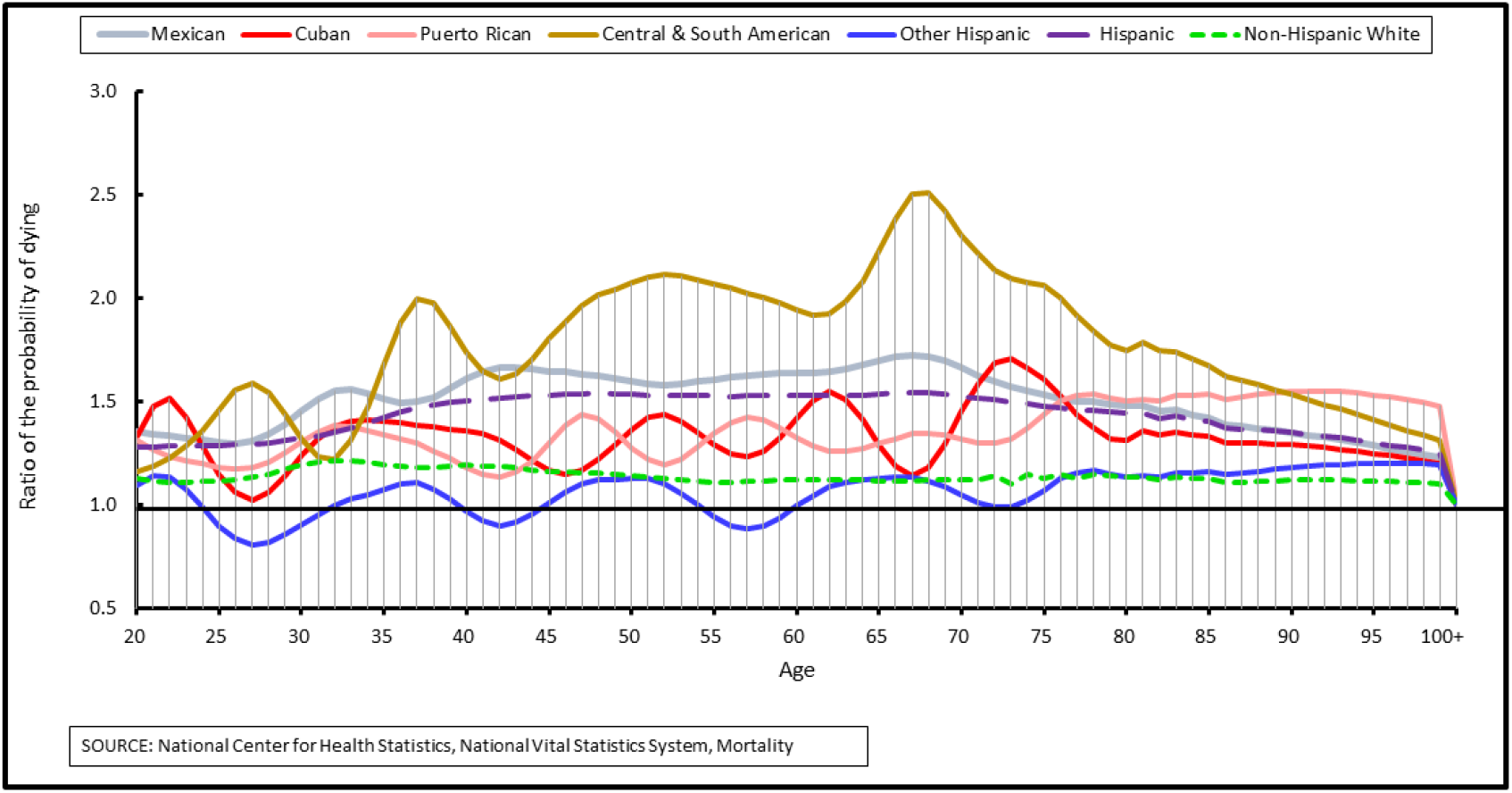
Age distribution of the relative increase in the probability of dying for males by Hispanic sub-group, 2019-2020.

**Figure 1b:**
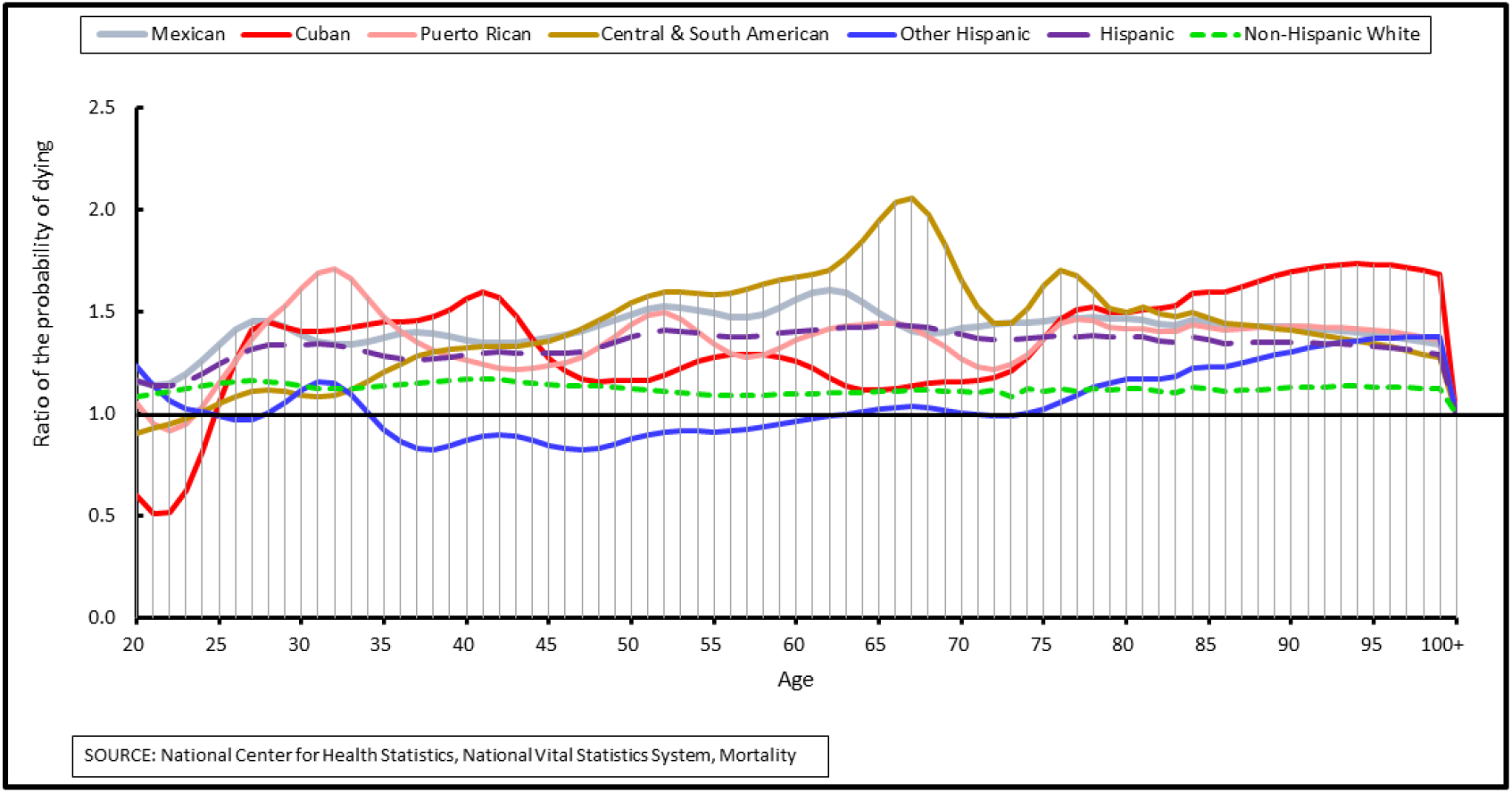
Age distribution of the relative increase in the probability of dying for females by Hispanic sub-group, 2019-2020.

There was considerable heterogeneity in the patterns and magnitude of the relative increases in age-specific mortality across Hispanic sub-groups. CSA males experienced the greatest increases in mortality throughout ages 35 to 85, peaking at age 68 with mortality in 2020 two and a half times greater than in 2019. Mexican males followed with increases in mortality between ages 30 and 70 that were larger than those of the other subgroups. The patterns of age-specific increases in mortality for Puerto Rican and Cuban males were similar and exhibited more variation. The age-specific pattern of mortality change for Other Hispanic males was relatively constant alternating with mortality increases and decreases of around 20% throughout most of the age-span. The much larger increases in mortality observed for Hispanic males appear to be driven mainly by CSA male followed by Mexican male mortality.

Among females, there was heterogeneity in the patterns and magnitude of age-specific changes in mortality, although not as large as for males. CSA females experienced the greatest increases in age-specific mortality in the middle to older ages, with a pronounced peak at age 67 where the probability of dying in 2020 was approximately two times greater than in 2019. Similar to their male counterparts, Mexican females followed with greater increases in mortality compared to the other Hispanic subgroups. Patterns for Cuban and Puerto Rican females were similar to each other with large increases in mortality in the younger adult ages. However, Cuban females experienced larger increases in mortality at ages over 80 years.

### Effects of changes in age and cause-specific mortality

Table 3 presents the results of the decomposition of the effects of changes in age-specific all-cause mortality to the decline in life expectancy between 2019 and 2020 (Arriaga 1984). For all groups, the percent contribution of mortality increases in childhood/adolescence was minimal. The contribution of increases in working-age adult mortality was greater for males than females across all groups. Conversely, the effects of increases in mortality at ages 70 and over were greater among females across all groups. The age-patterns of percent contributions to changes in life expectancy did not differ substantially between the overall Hispanic and non-Hispanic White populations, although the effects of increases in mortality for working age adults were greater for the non-Hispanic White than the Hispanic population.

**Table 3.** Percent contribution of increases in age-specific mortality to declines in life expectancy at birth between 2019 and 2020

|  | Males |  |  | Females |  |  |
| --- | --- | --- | --- | --- | --- | --- |
|  | Under 20 | 20-69 | 70 and older | Under 20 | 20-69 | 70 and older |
| Hispanic | 1.1 | 54.7 | 44.2 | 0.9 | 37.6 | 61.5 |
| Cuban | 2.0 | 39.5 | 58.4 | 0.7 | 17.9 | 81.4 |
| Mexican | 1.3 | 57.4 | 41.3 | 0.8 | 38.5 | 60.8 |
| Puerto Rican | 2.3 | 46.2 | 51.5 | 2.5 | 38.4 | 59.1 |
| Central & South American | 0.4 | 49.3 | 50.3 | 0.1 | 34.4 | 65.5 |
| Other Hispanic | 0.0 | 42.6 | 57.4 | 6.0 | 6.8 | 87.1 |
| Non-Hispanic white | 2.3 | 58.4 | 39.4 | 0.2 | 44.5 | 55.3 |

Notes: Other Hispanic includes Dominicans, Spaniards, and persons identifying as Hispanic/Latino, or Latin American.
Source: National Center for Health Statistics, National Vital Statistics System, Mortality.

There is heterogeneity within the Hispanic population in the effects of age-specific increases in all-cause mortality. Among males, the percent contribution of mortality increases for working age adults was greatest for Mexican (57.4%) and smallest for Cuban populations (39.5%). Conversely, the percent contribution of mortality increases for those aged 70 and older ranged from 41.3% for Mexican males to 58.4% for Cuban males. Among females, the percent contribution of mortality increases for working age adults were smaller for Other Hispanic and Cuban women, 6.8% and 17.9%, respectively, compared to the other Hispanic subgroups. The percentages for Mexican, Puerto Rican and CSA females were similar at 38.5%, 38.4%, and 34.4%, respectively. The largest effects of age-specific increases in mortality occurred in the oldest ages for Cuban and Other Hispanic females (81.4% and 87.1%, respectively). While still large, the contributions of increases in mortality in the oldest ages were lower for CSA, Mexican, and Puerto Rican females (65.5%, 60.8%, 59.1%, respectively).

Figure 2 presents the results of the decomposition of the effects of changes in cause-specific mortality to the decline in life expectancy between 2019 and 2020 (Arriaga, 1984). Increases in mortality due to COVID-19 had the greatest impact on declines in life expectancy in comparison to all other causes of death, for both the Hispanic and non-Hispanic White populations. However, COVID-19 had a larger effect on the Hispanic population. Increases in mortality due to COVID-19 explained 73.3% and 71% of the decline in life expectancy among Hispanic males and females, respectively. In contrast, the effects for non-Hispanic White males and females were 53.8% and 60.5%, respectively.

**Figure 2.**
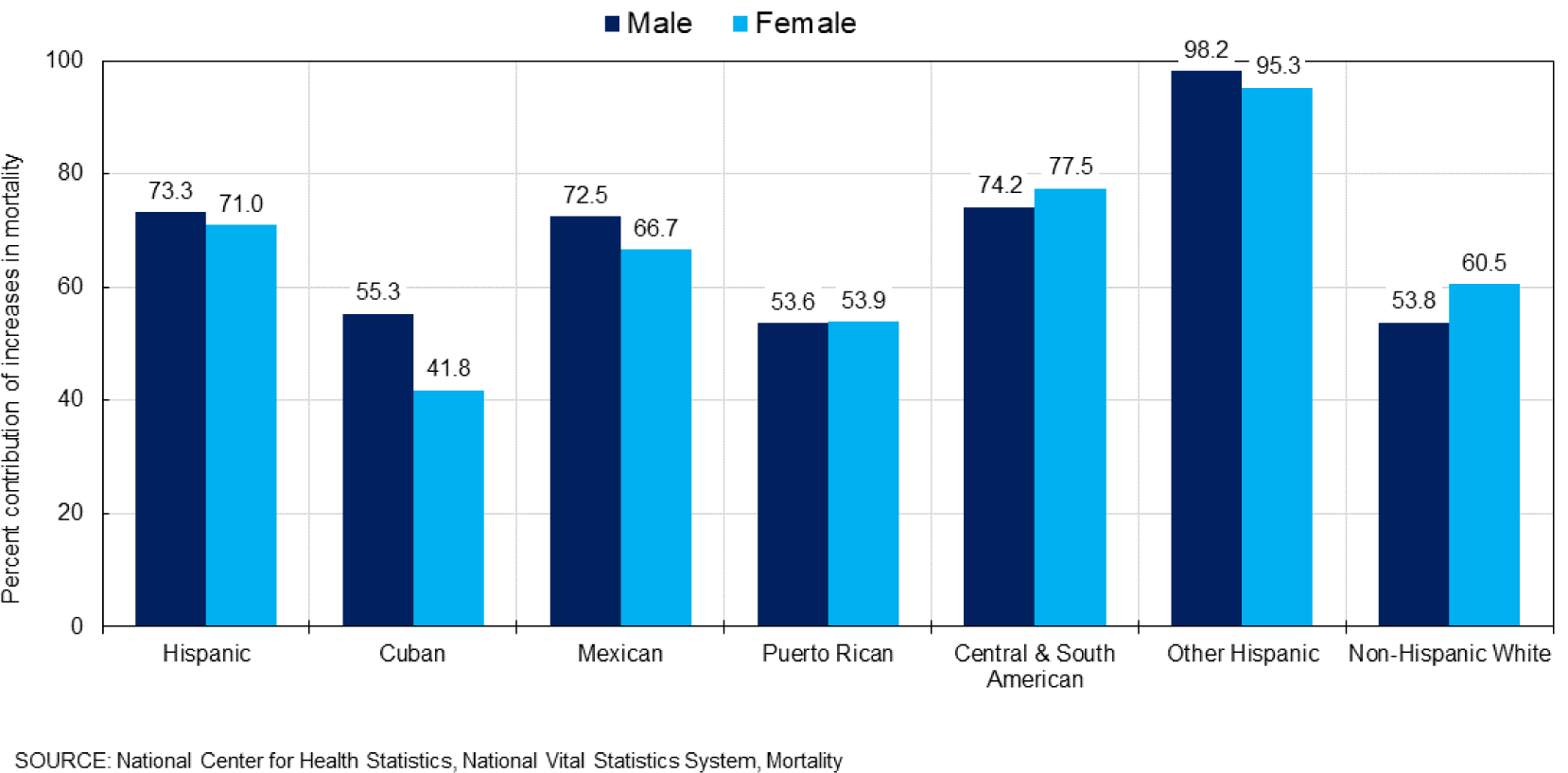
Percent contribution of increases in mortality due to COVID-19 to declines in life expectancy at birth between 2019 and 2020.

There was variation within the Hispanic population in the effects of increases in mortality due to COVID-19. The percent contribution of increases in mortality due to COVID-19 for males were smallest for Puerto Rican and Cuban at 53.6% and 55.3%, respectively, compared to 74.2% for CSA and 72.5% for Mexican males. The small decline in life expectancy (0.6 year) for Other Hispanic males was almost entirely explained by COVID-19 mortality (98.2%). Cuban females experienced the smallest effect of increases in COVID-19 mortality on change in life expectancy (41.8%) while the effects for CSA and Mexican females were larger at 77.5% and 66.7%, respectively. Like their male counterparts, COVID-19 explained almost all of the 0.6-year decline in life expectancy for Other Hispanic females (95.3%).

## Conclusions

The goal of this study was to explore how excess mortality due to the COVID-19 pandemic differentially affected Hispanic sub-groups. We estimated life expectancy at birth by sex and compared losses in life expectancy between 2019 and 2020 for each group. We explored how relative increases in age-specific mortality differed and assessed the direct effects of age- and cause-specific increases in mortality on the decline in life expectancy by sub-group.

Results show that life expectancy loss varied greatly by Hispanic sub-group and sex. CSA and Mexican males experienced declines in life expectancy that were more than two times greater than those of some of their other male counterparts. Group differences among females were smaller. Other Hispanic males and females experienced unusually small declines in life expectancy compared to the other Hispanic groups, but they had the lowest life expectancy and were the only sub-groups that experienced a mortality disadvantage relative to the non-Hispanic White population before the pandemic.

The impact and patterns of increases in all-cause age-specific and COVID-19 mortality on the decline in life expectancy also differed greatly across subgroups. CSA and Mexican males experienced greater increases in mortality overall and in the working ages than Cuban, Puerto Rican, or Other Hispanic males. The decomposition revealed that increases in mortality in working ages had the largest effect on the decline in life expectancy for Mexican and CSA males. CSA and Mexican females experienced larger increases in mortality in the working ages while Cuban and Puerto Rican females experienced greater increases in mortality at the oldest ages. The decomposition showed that increases in mortality in the oldest ages had the greatest effect on the decline in life expectancy for all Hispanic females but there were differences across the sub-groups. Increases in mortality in the oldest ages explained most of the decline in life expectancy among Cuban females but substantially less for other females. Finally, the impact of increases in mortality due to COVID-19 on the decline in life expectancy also differed across Hispanic sub-groups.

A previous study of the factors behind the disproportionate effect of the COVID-19 pandemic on Hispanic mortality posited that the excess burden for the Hispanic population can be explained by Hispanics’ overrepresentation in essential industries with higher risks of exposure to the virus (Do and Frank 2021). Our findings may possibly lend support to this hypothesis but only for some Hispanic sub-groups, like the CSA and Mexican populations, who experienced the greatest increases in mortality in the working ages and had greater proportions of the civilian employed population in occupations with greater risks of exposure to the virus (Billock, Steege and Minino 2022; Goldman et al. 2021). The same study reports that it found no support for the hypothesis of intergenerational transmission of the virus in multigenerational Hispanic households because excess mortality in the oldest ages was relatively low in the Hispanic population (Do and Frank 2021). Our results suggest that this may be the case for some Hispanic sub-groups. We found that for Cuban females, the contribution of COVID-19 mortality to the decline in life expectancy was concentrated in the oldest ages and therefore could possibly be a function of the greater prevalence of multigenerational families in the Cuban population, for example (Palloni and Arias 2004).

This study has important limitations. First, our life expectancy estimates are based on mortality data that have been adjusted for Hispanic origin misclassification on death certificates and statistical modeling. As a result, the estimates are not free from error. Second, the Other Hispanic and CSA subgroups combine diverse populations that likely have distinct mortality profiles. To test the validity of our sub-group life expectancy estimates, we estimated the life expectancy of the entire Hispanic population as the population weighted sum of the individual sub-group life expectancies. Ideally, our estimate should be very close to the official NCHS estimate. For 2019, our estimate was 0.2 year lower than the official estimate (81.7 vs. 81.9) and for 2020 it was 0.5 year lower (77.4 vs. 77.9). Despite these limitations, our study underscores the importance of population disaggregation in analyses of the multiple pathways by which the pandemic so disproportionately affected the Hispanic population.

## Data Availability

All data produced in the present work are contained in the manuscript. Mortality data source used to produce estimates contained in the manuscript can be accessed at: https://www.cdc.gov/nchs/nvss/deaths.htm. Population data source used to produce estimates contained in the manuscript can be accessed at: U.S. Census Bureau, 2020 American Community Survey 1-Year Estimates. Available from: https://www.census.gov/programs-surveys/acs/data.html. U.S. Census Bureau, 2019 American Community Survey 1-Year Estimates. Available from: https://www.census.gov/programs-surveys/acs/data.html.

## References

Andrasfay, T, and Goldman, N. (2021). Reductions in 2020 US life expectancy due to COVID-19 and the disproportionate impact on the black and Latino populations. Proceedings of the National Academies of Science 118(5): 1–6.

Arias, E, and Xu J. (2022). United States life tables, 2020. National Vital Statistics Reports, 71(1). National Center for Health Statistics.

Arias, E, Tejada-Vera, B, Kochanek, KD, and Ahmad, FB. (2022). Provisional life expectancy estimates for 2021. Vital Statistics Rapid Release Report No. 23. National Center for Health Statistics.

Arias, E, Heron, M, Hakes, JK. (2016). The validity of race and Hispanic origin reporting on death certificates in the United States: An update. Vital and Health Statistics Series Report 2 (172). National Center for Health Statistics.

Arriaga, EE. (1984). Measuring and explaining the change in life expectancies. Demography 21(1): 83–96.

Do, DP, and Frank, R. (2021). Using race- and age-specific COVID-19 case data to investigate the determinants of the excess COVID-19 mortality burden among Hispanic Americans. Demographic Research 44(29): 699–718.

Billock, RM, Steege, AL, Miniño, A. (2022). COVID-19 mortality by usual occupation and industry: 46 states and New York City, United States, 2020. National Vital Statistics Reports 71 (6). Hyattsville, MD: National Center for Health Statistics (forthcoming).

Fujishiro, K, Xu, J, Gong, F. (2010). What does “occupation” represent as an indicator of socioeconomic status?: exploring occupational prestige and health. Social Science and Medicine 71 (12): 2100–7.

Garcia, MA, Homan, PA, Garcia, C, and Brown, TH. (2020). The color of COVID-19: Structural racism and the disproportionate impact of the pandemic on older black and Latinx adults. Journal of Gerontology: Social Sciences 76(3): e75–e80.

Goldman, N, Pebley, AR, Keunbok, L, Andrasfay, T, Pratt, B. (2021). Racial and ethnic differentials in COVID-19-related job exposures by occupational standing in the US. PLOS One September 1, 2021: 1–17.

Macias Gil, R. Marcelin, JR, Zuniga-Blanco, B, Marquez, C, Mathew, T, Piggott, DA. (2022). COVID-19 pandemic: Disparate health impact on the Hispanic/Latinx population in the United States. The Journal of Infectious Diseases Perspective. 222: 1592–1595.

Markides, KS, and Coreil, J. (1986). The health of Hispanics in the southwestern United States: An epidemiologic paradox. Public Health Reports 101(3): 253–265.

Markides, KS and Eschbach K. (2011). Hispanic paradox in adult mortality in the United States. In R. Rogers and E. Crimmins (Eds.), International handbook of adult mortality. International handbooks of population, Vol. 2. Dordrecht: Springer.

National Center for Health Statistics. 2020. Vital statistics data available online: Mortality public-use file, 2019. Hyattsville, MD: National Center for Health Statistics. Published annually. Available from: https://www.cdc.gov/nchs/data_access/VitalStatsOnline.htm.

National Center for Health Statistics. 2021. Vital statistics data available online: Mortality public-use file, 2020. Hyattsville, MD: National Center for Health Statistics. Published annually. Available from: https://www.cdc.gov/nchs/data_access/VitalStatsOnline.htm.

Palloni, A, Arias, E. (2004). Paradox lost: Explaining the Hispanic adult mortality advantage. Demography 41(3): 385–415.

Riley, AR, Chen, YH, Matthay, EC, Glymour, MM, Torres, JM, Fernandez, A, and Bibbins-Domingo, K. (2021). Excess mortality among Latino people in California during the COVID-19 pandemic. SSM – Population Health. 15 (2021) 100860.

Saenz, R. and Garcia, MA. (2020). The disproportionate impact of COVID-19 on older Latino mortality: The rapidly diminishing Latino paradox. Journal of Gerontology: Social Sciences 76(3): e81–e87.

U.S. Census Bureau, 2020 American Community Survey 1-Year Estimates. Available from: https://www.census.gov/programs-surveys/acs/data.html.

U.S. Census Bureau, 2019 American Community Survey 1-Year Estimates. Available from: https://www.census.gov/programs-surveys/acs/data.html.

